# Distribution of the 2019-nCoV Epidemic and Correlation with Population Emigration from Wuhan, China

**DOI:** 10.1101/2020.02.10.20021824

**Authors:** Zeliang Chen, Qi Zhang, Yi Lu, Zhongmin Guo, Xi Zhang, Wenjun Zhang, Cheng Guo, Conghui Liao, Qianlin Li, Xiaohu Han, Jiahai Lu

## Abstract

**BACKGROUNDS:** The ongoing new coronavirus (2019-nCoV) pneumonia outbreak is spreading in China and has not reached its peak. Five millions of people had emigrated from Wuhan before the city lockdown, which potentially represent a source of virus spreaders. Case distribution and its correlation with population emigration from Wuhan in early epidemic are of great importance for early warning and prevention of future outbreak.

**METHODS:** The officially reported cases of 2019-nCoV pneumonia were collected as of January 30, 2020. Time and location information of these cases were extracted analyzed with ArcGIS and WinBUGS. Population migration data of Wuhan City and Hubei province were extracted from Baidu Qianxi and analyzed for their correlation with case number.

**FINDINGS:** The 2019-nCoV pneumonia cases were predominantly distributed in Hubei and other provinces of South China. Hot spot provinces included Sichuan and Yunnan provinces that are adjacent to Hubei. While Wuhan city has the highest number of cases, the time risk is relatively stable. Numbers of cases in some cities are relatively low, but the time risks are continuously rising. The case numbers of different provinces and cities of Hubei province were highly correlated with the emigrated populations from Wuhan. Lockdown of 19 cities of Hubei province, and implementation of nationwide control measures efficiently prevented the exponential growth of case number.

**INTERPRETATION:** Population emigrated from Wuhan was the main infection source for other cities and provinces. Some cities with low case number but were in rapid increase. Due to the upcoming Spring Festival return transport wave, understanding of the trends of risks in different regions is of great significance for preparedness for both individuals and institutions.

**FUNDINGS:** National Key Research and Development Program of China, National Major Project for Control and Prevention of Infectious Disease in China, State Key Program of National Natural Science of China.

## Introduction

Emerging infectious diseases are still great challenges in the 21st century. In recent years, worldwide outbreaks of Ebola and Middle East Respiratory Syndrome (MERS) caused great health and economic losses^1-3^. The ongoing pneumonia outbreak caused by the new coronavirus, 2019-nCoV, is becoming a global public health problem. The 2019-nCoV outbreak is highly similar to the Severe Acute Respiratory Syndrome (SARS) in 2003 that they were both caused by new coronaviruses and happened in time periods including Chinese Spring Festival^1^. On December 31, 2019, the Wuhan Municipal Health and Health Committee reported 27 cases of pneumonia with an unknown cause, and many cases were related to the Wuhan Southern China Seafood Market, which was subsequently closed on January 1, 2020^4^. On January 7, 2020, laboratory test results showed that the pathogen of unexplained pneumonia was a new type of coronavirus, which was then officially named 2019-nCoV by the World Health Organization (WHO)^5,6^. The 2019-nCoV outbreak started in Wuhan, and then spread rapidly to other provinces and countries^7,8^. As of January 30, 2020, a total of 34 provinces and regions in China have reported 9,692 cases. Nearly all imported cases were derived from Wuhan, Hubei province^9,10^.

The 2019-nCoV pneumonia was defined as a class B infectious disease but managed as class A infectious disease by the Chinese government. Daily case reports are being released and any omission or concealment is punishable by law. At present, the number of cases is still increasing and the epidemic has not yet reached its peak, but the situation differs by province.. Information about the temporal and spatial distributions of the cases is important for preparation of a targeted treatment and prevention strategy. Because the return peak of Spring Festival transport is coming, the information on possible changes in incidence in different cities will help us to prepare in advance. Therefore, we investigated the temporal and spatial distributions of the early epidemic to reveal the dynamic changes and trends. These results will provide valuable information for the disease prevention for both individuals and organizations.

## Methods

### Case data collection

All officially reported confirmed, suspected, severe cases and deaths of 2019-nCoV pneumonia were collected from the internet (website:). Case data were imported into Excel and incidence per 100,000 was calculated. No ethical approval is needed for this study.

### Temporal and spatial distribution and risk analysis

The national and Hubei province shapefiles are used for ArcGIS analysis. The map is linked to Excel file with time and spatial information. The risk analysis of the 2019-nCoV pneumonia is based on the Bayesian space-time model of WinBUGS software^11,12^. The model is divided into three levels:

#### (1) Data model

For the statistical data of low incidence, it is assumed to follow the Poisson distribution of the parameters n_i_ and μ_it_: y_it_ ∼ Poiss (n_i_μ_it_), where Hubei province y_it_ is i (1,…., 16) cities at t (1,…, 11) the number of cases that occurred during the day. The nationwide y_it_ is the number of cases that occurred in t (1,…, 11) days in i (1,…., 240) cities. We assume that there is no change in the number of people at risk in each city during the study period, where n_i_ is the number of people at risk in the town (i), and μ_it_ is the corresponding disease risk in the city (t) per day (i).

#### (2) Process model

The μ_it_’s logarithmic transformation of disease risk allows relative risk to be expressed as a linear combination of a spatial component, a temporal component, and a spatiotemporal interaction component. The mathematical expression is shown in equation (2).

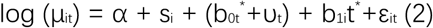

Where α is the fixed effect of the overall relative risk of the entire study area within 11 days; t^*^ = t-5.5 is the time span relative to the intermediate time point. In this model, the risk of disease is broken down into three parts: spatial change, temporal change, and space-time interaction; and s_i_ is a component of spatial variability, describing the urban disease risk relative to the entire study region over an 11-day observation period; b_0t_^*^+υ_t_ is the amount of change over time, which describes the overall trend of disease risk in the entire study area relative to the medium-term observation day, including the linear trend b_0t_^*^ and the time random effect υ_t_, b_0_ is the time coefficient, represents the time trend of the study area; b_1i_t^*^ allows each city to have different time-varying trends and is part of the spatiotemporal interaction. Relative to b_0_, it is the local change trend of each city based on b_0_; ε_it_ is used to explain local changes that cannot be explained by spatiotemporal random effects.^13^

#### (3) Parametric model

According to the BYM model^14^, a spatial structure effect is defined by a prior conditional autoregressive (CAR) structure. In this process, a spatial adjacency weight matrix needs to be defined. The weight adjacency is w_ij_=1, otherwise the weight w_ij_=0, and the special wij=0. Similarly, b_1i_ is also assumed to follow the BYM process. For the time structure effect υ_t_, a conditional autoregressive process is used, and the adjacency weight matrix in time is defined. For the over-discrete parameter ε_it_, according to Gelman, the normal distribution with a mean value of 0 and a variance of σ^2^ε, is generally assumed and the variance of each parameter obeys Gamma (a, b).^15^ Based on this model, through the spatial component s_i_ and its posterior probability, high-risk or low-risk cities with respect to the average risk (α) of the entire study area can be identified. By calculating the probability of spatial relative risk, exp (s_i_) is greater than 1 and divided into five categories: areas with probability> 0.8, 0.6-0.8, 0.4-0.6, 0.2-0.4, and <0.2 are defined as hot spots, secondary hot spots, warm-spots, sub-cold spots, and cold-spots respectively. Similarly, based on the probability threshold, the differences in these classification regions can be identified based on the trend over time. The probability that exp (b_1i_) is greater than 1 is divided into five categories: cities with an incidence risk probability greater than 0.8 have a rapid change trend relative to the overall risk change; between 0.6 and 0.8 indicate that the change trend of the incidence risk is greater than the overall risk change; 0.4 to 0.6 indicates that the occurrence risk is the same as the overall risk change trend; 0.2 to 0.4 indicates that the change trend of the disease risk is slower than the overall risk change; the change trend of the disease risk in cities with a probability less than 0.2 is more slower than the overall risk.

### Correlation between case number and population migration

Population migration data were collected from Baidu dataset (http://qianxi.baidu.com/). Data on emigration from Wuhan City and Hubei province to other cities and provinces were extracted and edited with Excel of windows Microsoft office. Emigration intensity was calculated as migration index multiplied with proportions of the province or city.

## Results

To obtain a general profile of the distribution, we first analyzed all the available cases during this 2019-nCoV pneumonia outbreak^16^. As shown in Figure 1A, the number of cases remained stable from January 11 to 15, 2020, and the number of newly added and cumulative cases increased rapidly after January 16. The first death was reported on January 10, and the number of deaths began to increase rapidly from January 17, with the cumulative death number reaching 312 on January 30 (Figure 1B)^6^. After the nucleic acid assay becoming available, suspected cases waiting for laboratory confirmation could now be diagnosed more rapidly ^17^. After January 19, suspected cases increased rapidly, and about 40-50% of these suspected cases were then diagnosed as confirmed cases (Figure 1C). Before January 19, the number of severe cases, remained at a low level but increased steadily since January 20 (Figure 1D). Because Wuhan is the capital city of Hubei province and the cases have spread to the rest of Hubei province quickly, we also analyzed the change in number of cases in Hubei province. On January 9, 41 cases were first reported, and by January 30, 5,806 cases were reported, accounting for 59.91% of the total cases in China (Figure 1E). The cumulative number of deaths in Hubei province is 249, accounting for 96.14% of total deaths in China (Figure 1F). These data indicated that both the incidence and mortality of 2019-nCoV disease is the highest in Hubei province^18^.

**Figure 1.**
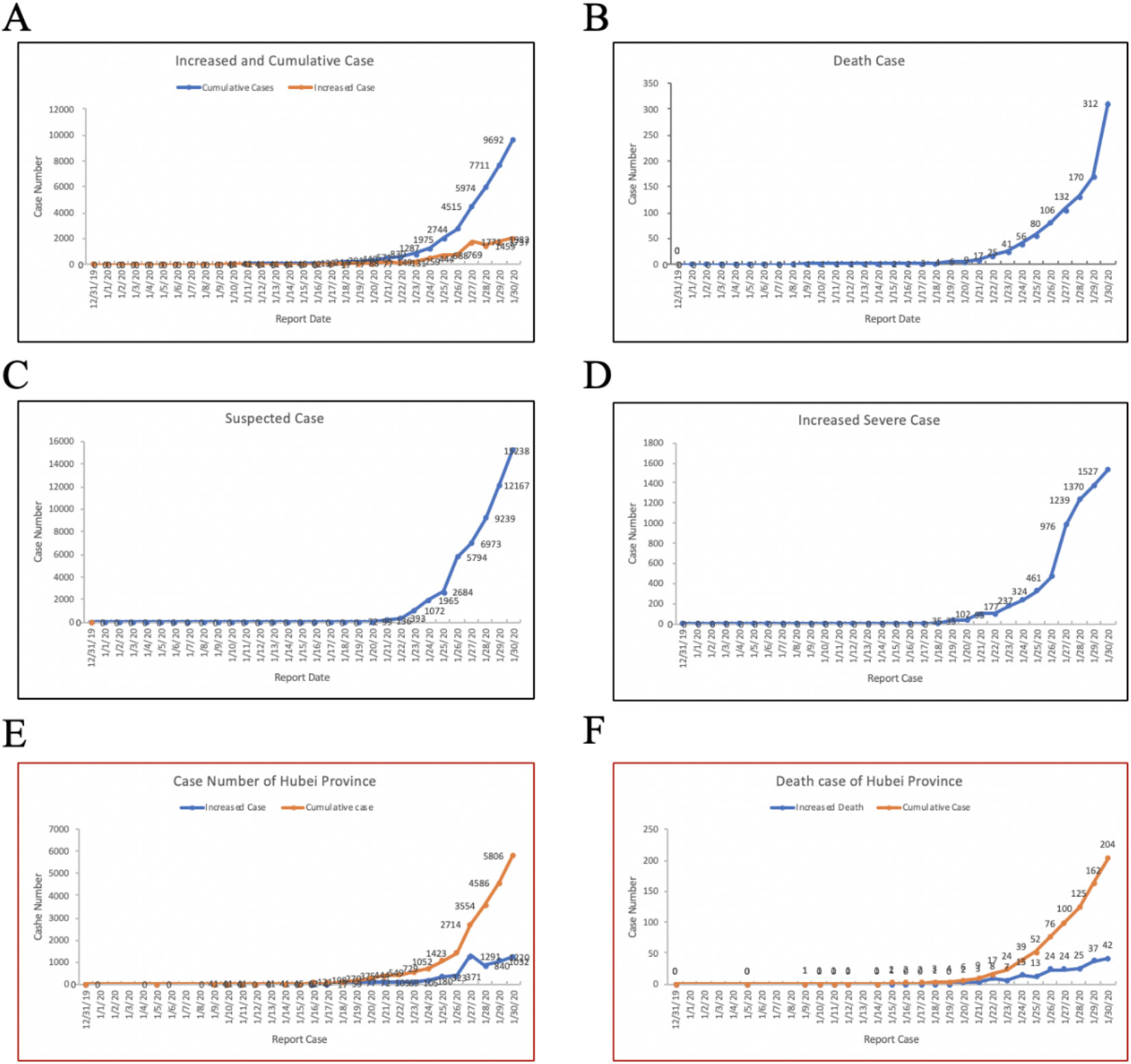
2019-nCoV Case Report of China and Hubei Province. A-D, Daily report on national increased and cumulative confirmed cases (A), death case (B), suspected cases (C), and increased severe cases (D). E, Increased and cumulative case number of Hubei Province; D, Increased and cumulative death case number of Hubei province. A, Daily increased and cumulative case number.

Before January 16, cases were mainly reported in Hubei province. From January 17 onward, the outbreak spread to many provinces and the number of cases increased rapidly. Therefore, our spatial and temporal analyses used data from January 17 to January 30, 2020. Location of each case was extracted from official reports and mapped on the national map to city level with ArcGIS. Of the 362 cities, 307 (84.8%) had reported cases. In general, the core outbreak area, Wuhan and its surrounding cities, had the highest number of cases, followed by cities with high population that are transportation hubs (Figure 2A). Spatial distribution was then analyzed with a Bayesian model using WinBUGS^13,14^. After iterating nearly 100,000 times, the model converged successfully. After the model converged, it iterated another 110,000 times to obtain parameter estimations. A ratio close to 1 indicates that the two chain iterative sequences are close, and that the model has a good convergence and is stable (Figure 2B). With the established model and parameters, hot and cold spots were identified. The results showed that Sichuan, Yunnan, Guizhou, Hainan, and Taiwan were hot spots, and Inner Mongolia, Gansu, Ningxia, Qinghai, Xinjiang, Chongqing, Hunan, and Guangxi were secondary hot spots. Generally, hot spots clustered in the Midwest and cold spots clustered in the Southeast (Figure 2D).

**Figure 2.**
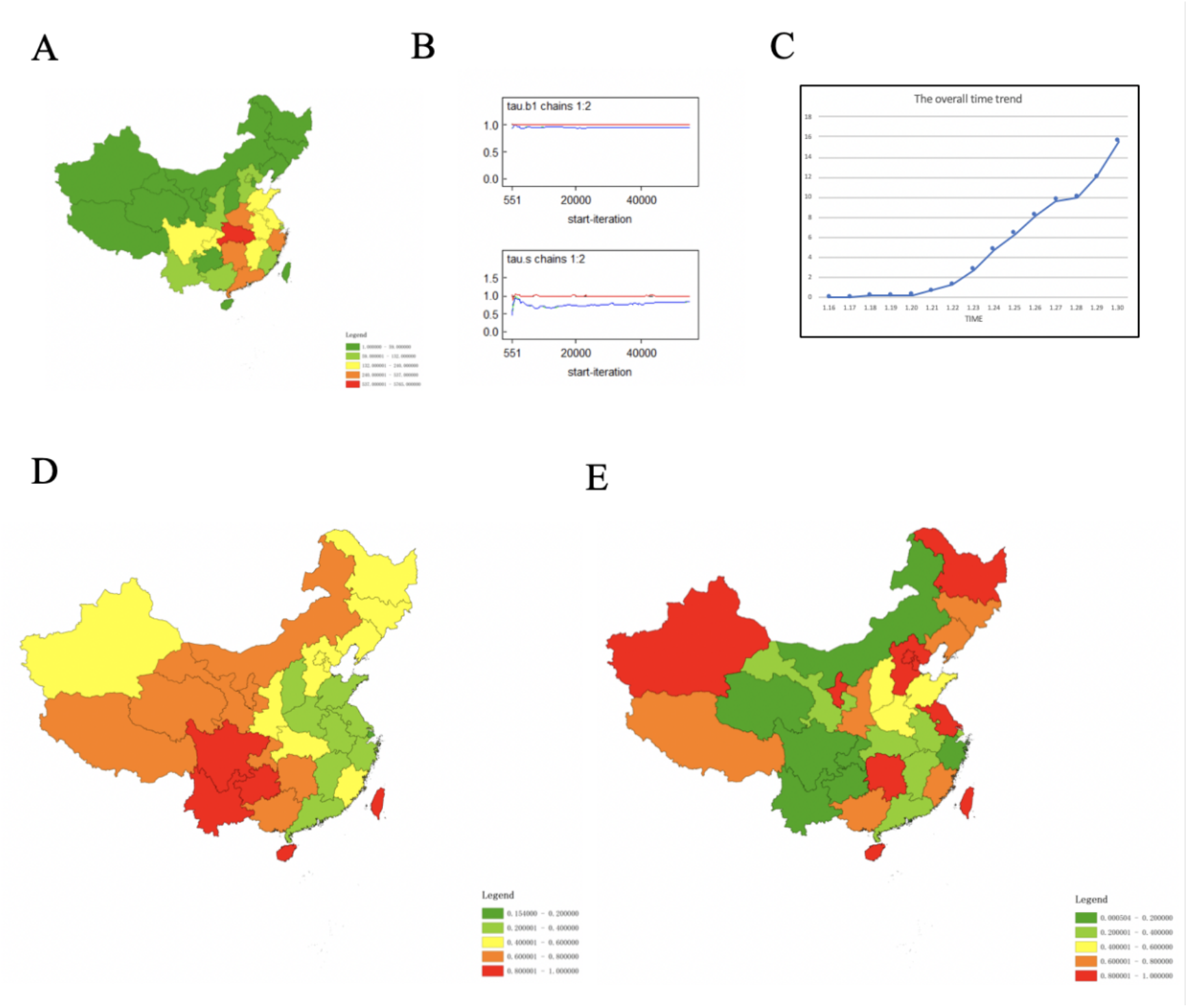
Nationwide Case distribution and change trends. A, Case distribution, B, Hot and cold spot of case distribution; C, model convergence analysis; D, Changing trends of case distribution.

The overall temporal trend was calculated using the time risk model (exp(b_0t_*+v_t_)), which described the general incidence risk by time during the period from January 16 to January 30, 2020. The results showed that b_0_ is estimated to be 0.4604, that is, the disease risk on the following day is approximately 1.585 times higher than the previous day. The relative risk by time increased steadily from January 20 and remained in an upward trend as of January 30 (Figure 2C). From this, it can be estimated that the number of cases nationwide is on the rise. As shown in Figure 2E, Heilongjiang, Hebei, Beijing, Tianjin, Xinjiang, Ningxia, Jiangsu, Hunan, Taiwan, and Hainan showed faster increased in cases than the overall national trend. The increase of cases in Jilin, Liaoning, Shanxi, Guangxi, and Fujian provinces were also relatively fast. The increase of cases in other provinces were consistent with or lower than the overall national trend.

Since Hubei province had the highest number of cases, we analyzed the temporal and spatial distribution in different cities of Hubei province. According to the reported cases, Wuhan had the highest number of cases, followed by Huanggang and Xiaogan city. Suizhou, Jingmen, and Xianning city are among the second group with a high number of cases (Figure 3A). The spatial convergence analysis converged 100,000 iterations (Figure 3B). Hot spots are identified in the east regions and cold spots are in the west regions (Figure 3D). The overall temporal trend in the model estimation was calculated for the change in the number of cases. The average time trend coefficient b_0_ was estimated to be 0.6727, meaning the time risk (occurrence probability in time) on a following day is 1.960 times higher than the previous day, indicating that the daily number of cases in Hubei Province is on the rise (Figure 3C). Xiangyang, Suizhou, Yichang, and Ezhou showed the highest increase rates, and Shiyan, Shenlongjia, Xiaogan and Huangshi showed relative high increase rates (Figure 3E). Other cities had a growth slower than the overall growth in the province (Table S1). The increase rate of Hubei province (1.960) was higher than the whole country (1.585), indicating the increase rate in Hubei province is significantly higher than other provinces in China.

**Figure 3.**
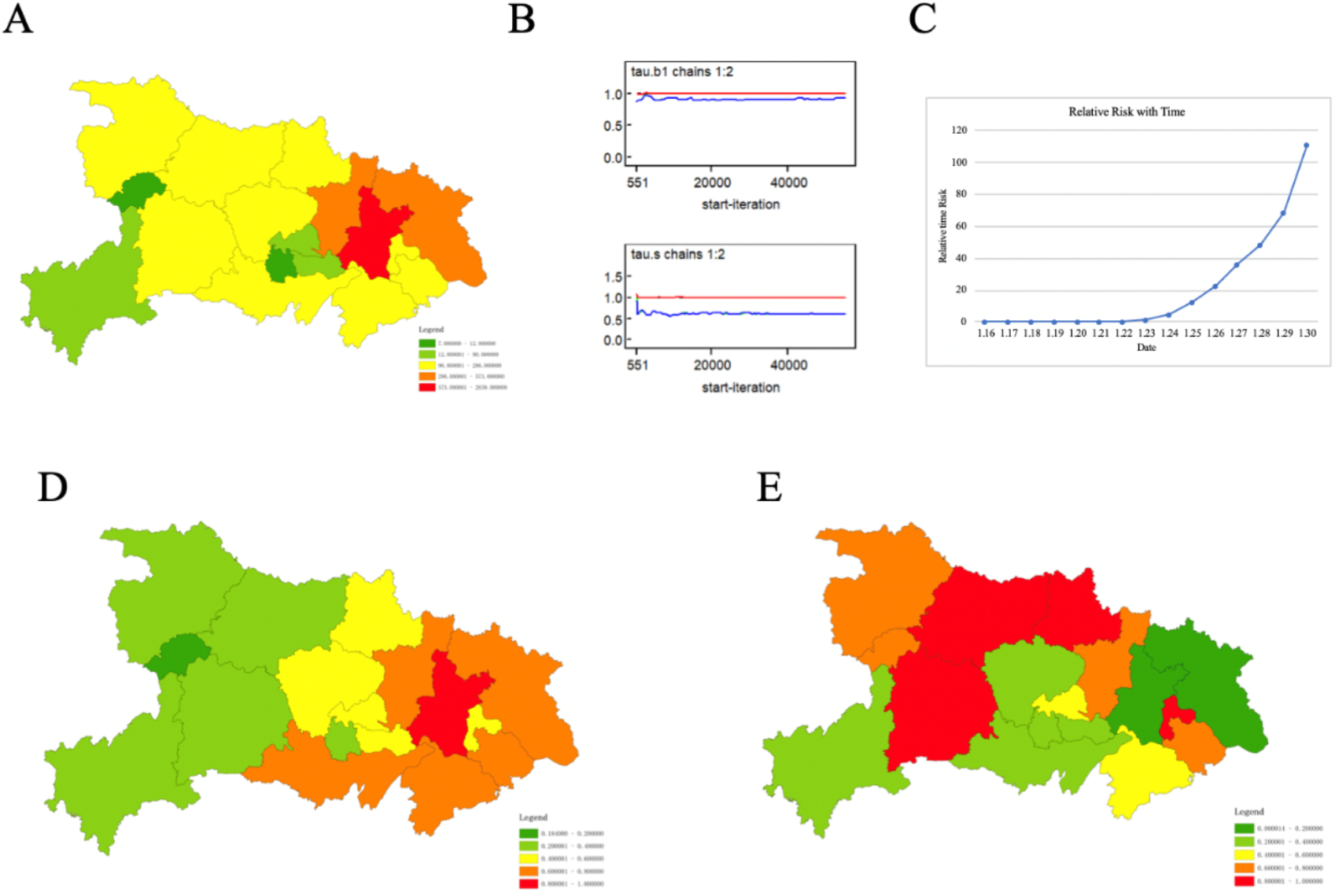
2019-nCoV Case distribution of Hubei province. A, Case distribution of Hubei province; B, Model convergence case distribution; C, Relative time risk with time; D, Hot spot and cold spot of case distribution; E, Incidence changing trends of different cities of Hubei Province.

In addition to changes in number of cases, we also analyzed the change in incidence, which could incorporate the population information of each city. Consistent with the trend of number of cases, Wuhan had the highest incidence, followed by Xiaogan and Huanggang (Figure 4A). Model converged after 110,000 iterations (Figure 4B). Spatial distribution trend analysis showed that Jingmen, Xiaogan, Qianjiang, Xiantao, Ezhou, and Huangshi were hot spots with high incidence, and Huanggang, Wuhan, Tianmen, Jingzhou, Xianning were cold spot with relatively low incidence (Figure 4C). Temporal change trend analysis results that the average time trend coefficient b_0_ was 0.5087, and the disease risk on the following day was approximately 1.663 times higher than the previous day, indicating an upward trend (Figure 4D). The change in incidence is different from the change in number of cases.

**Figure 4.**
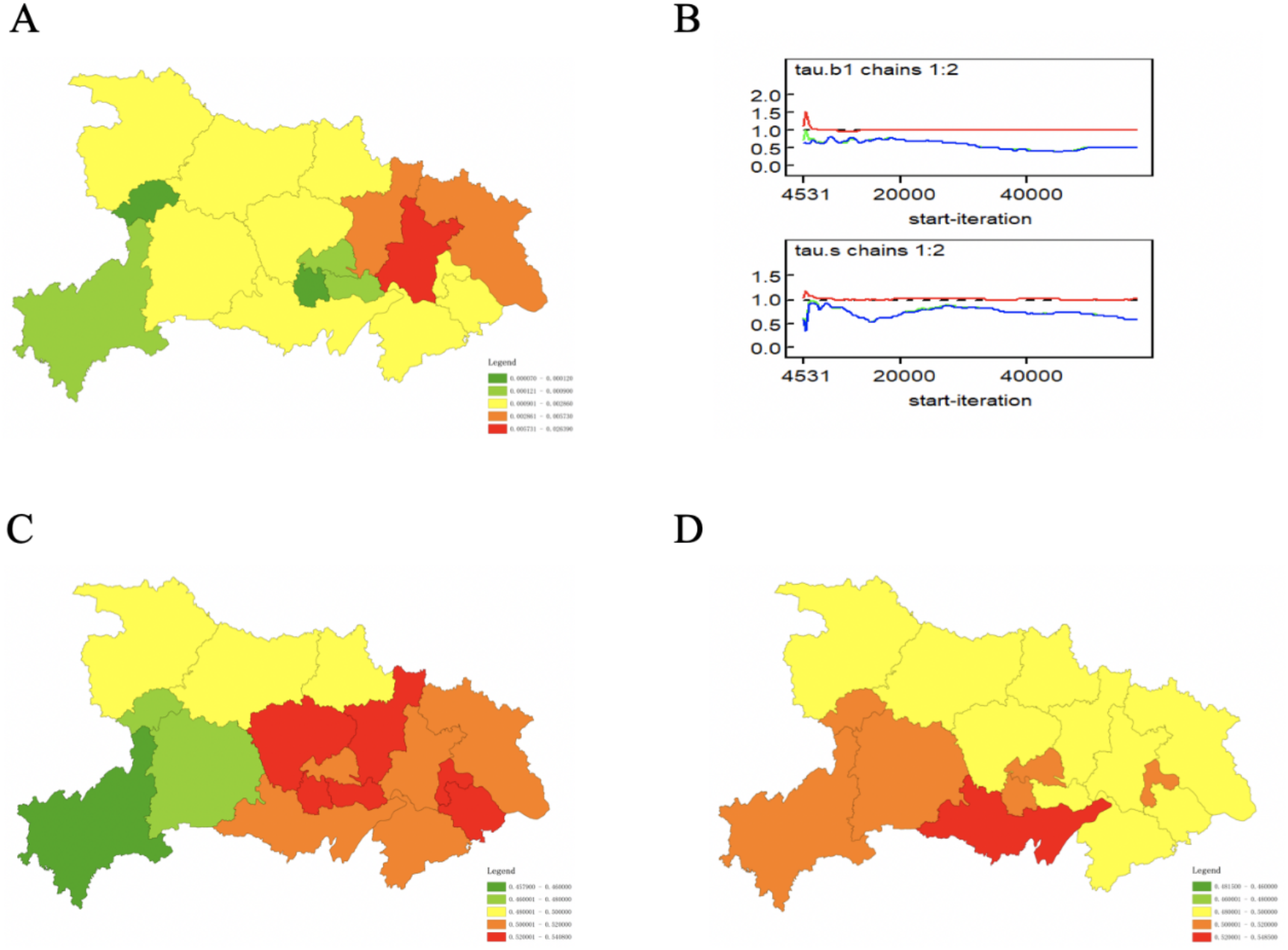
2019-nCoV incidence distribution and change trends of Hubei province. A, Incidence distribution of Hubei province; B, Model convergence of incidence distribution; C, Hot spot and cold spot of incidence distribution; D, Incidence changing trends of different cities of Hubei Province.

The outbreak started from Wuhan, and nearly all the early cases were derived from Wuhan, Hubei province. Because the outbreak happened just before Spring Festival, large scale population migration during this period is an inducing factor for subsequent epidemic. From Jan 1 to 23, 2020, the population migrated out of Wuhan city and Hubei province increase steadily and peaked at Jan 22 and 21 respectively (Figure 5A). Wuhan city is lockdown on Jan 23, and after that population migration greatly inhibited. As experienced 2019, high population migration occurred Jan 31, this timely city lockdown prevented subsequent burst. We then analyzed the migration out and into Wuhan city and Hubei province. The top target provinces of emigration included Henan and Hunan province (Figure 5B). More people migrated out of Wuhan than those into the city (Figure 5C). To analyze the correlation between case number and the emigration population for Wuhan city and Hubei province, population migration data was collected from Baidu Migration. The correlation coefficient between provincial case number and emigration of Hubei province was up to 0.821 (Figure 5D). The correlation coefficient between provincial case number and emigration from Wuhan was increased to 0.8895, and the highest one was 0.9918 between Wuhan and other cities of Hubei provinces (Figure 5E, F, Table S3, S4). These data strongly indicated that the case number is highly related with population emigration from Wuhan. Although we do not know the exact population emigrated from Wuhan, 5 million was an astonishing number, for each of them might be a virus spreader. If no control measures implemented, case number would exponentially increase. Of the 5 million emigrants, 74.22% were emigrated to other cities of Hubei province (Figure S2, Table S3). Fortunately, 19 cities of Hubei province were lockdown from Jan 23 to 26 (Figure S5). After lockdown of Wuhan and other cities of Hubei province, the outbreak burst was prevented, and case number increase only steadily without exponential growth.

**Figure 5.**
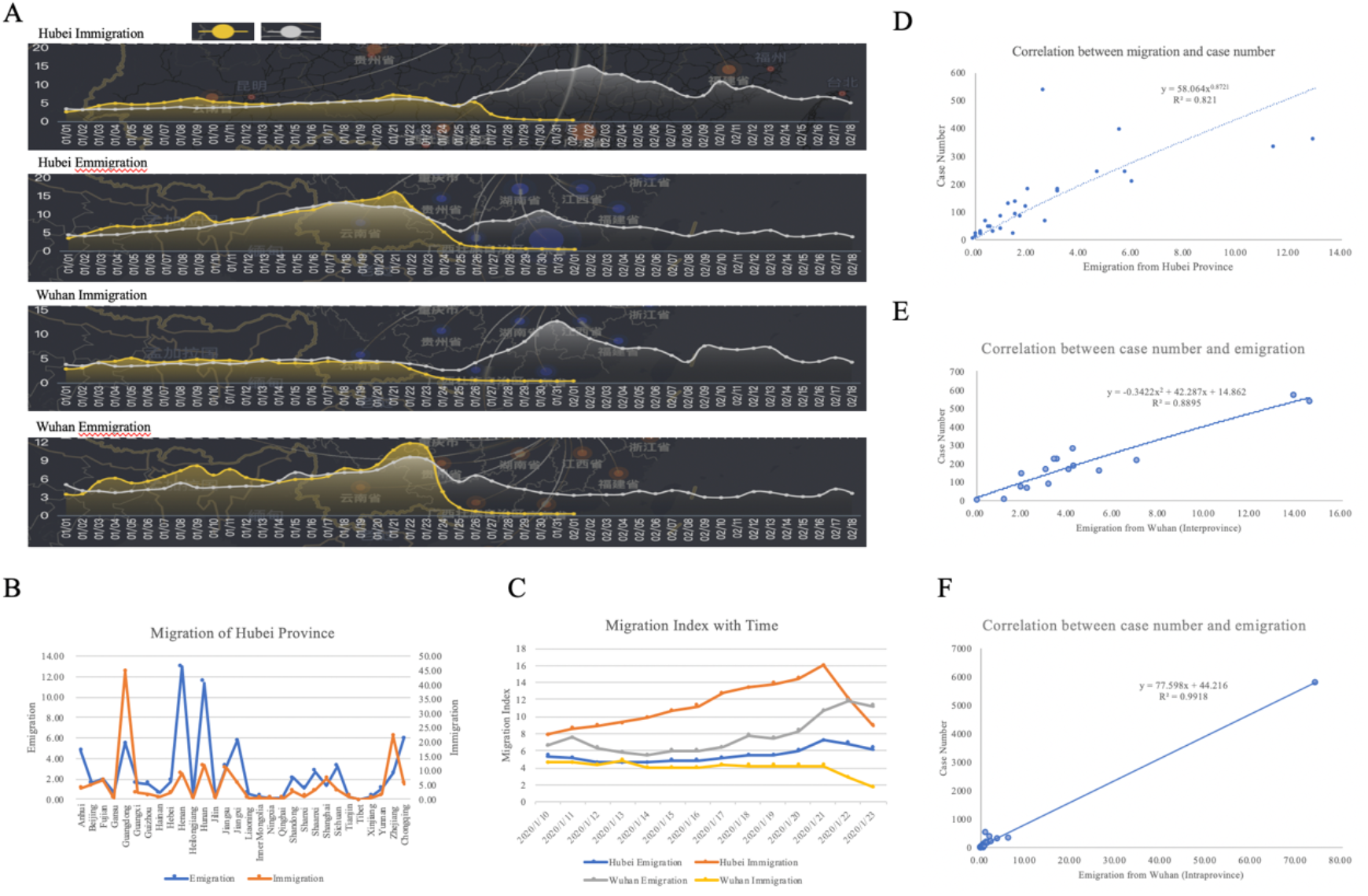
Correlation between Migration and Case Number. A, Migration index from and into Wuhan city and Hubei province during spring festival transport. Yellow, 2020, Gray, 2019. B, Migration out (emigration) and into Hubei Province; C, Emigration and immigration of Wuhan City and Hubei province from Jan 10 to 23, 2020; D, Correlation between case number and emigration from Hubei province; E, Correlation between case number and emigration from Wuhan city (interprovince); F, Correlation between case number and emigration from Wuhan city (intraprovince).

In on our previous trend analysis of the early spread of this outbreak and the previous SARS outbreak, we estimated that the cumulative number of 2019-nCoV cases might be 2-3 times the cumulative number of SARS cases and reaches between 16,000 and 24,000 cases.^19^ Because the outbreak duration overlaps the Spring Festival transport waves, large scale migration will be a decisive factor of this outbreak. We analyzed the migration in the 3 days before the Spring Festival. The top 50 cities that were exporting migrants before Spring Festival are mainly located in the south and east of China. The top 4 exporting cities include Beijing, Shenzhen, Shanghai, and Guangzhou, which account for over 15% of the migration population (Figure S1). However, the cities with high population of imported migrants were relatively scattered. Chongqing had the highest population of imported migrants, which accounted for 1.50% of the total imported migrants (Figure 5). As the migrants will be traveling back to work after the Spring Festival, these previous “exporting” cities may have a high risk of emergence of another wave of new cases caused by the returned migrants.

## Discussion

The 2019-nCoV pneumonia are causing great public health and economic losses for China. The case number increased rapidly, with over 50% were from Hubei province. As of Jan 30, the case number has exceeded the total number of the SARS-CoV outbreak^20^. To cope with the outbreak, the Spring Festival holiday had been extended for two times. Prevention and control of the outbreak became a concerted action of the whole people in China. Although all people participated in the campaign against the outbreak, people in area with low case number considered that the disease is far away. Therefore, awareness of high-risk regions is import for preparation and responses for all the people, particularly those in regions with low incidence. Another reason we need to keep in mind is that 5 millions of persons have been emigrated from Wuhan to all over the country, we do not know exact who of them are virus carriers, and it is impossible to track and diagnose them all. Therefore, isolation at home and less contact with others is the most efficient measure to prevent infection and transmission.

With the reported cases, we analyzed the temporal and spatial distribution. In general, the number of cases is still on the rise. For Hubei Province, which has the highest number of cases and deaths, the growing trend is relatively stable. Conversely in other hot spots, the number of cases were not so high, but the rate of growth continues to rise and therefore should be closely monitored. It is particularly noteworthy that cities with the fastest change in temporal risk, such as Chongqing, have large population movements and rapid temporal risk. If they are not strictly monitored, there may be more outbreaks. In order to prevent disease outbreaks caused by the returning travel wave to the city after the Spring Festival, the country has extended the Spring Festival holiday twice to February 9, 2020. However, as the number of new cases is still increasing, the outbreak is unlikely to end before the end of the holiday. Therefore, we must act together to stop the spread of the disease. At present, the state has adopted mobility control measures to encourage people to avoid going to public places and wear masks when going out to reduce the risk of human to human transmission. We believe that with the joint efforts of everyone, the number of cases and losses will be kept to a minimum.

## Data Availability

Data available from corresponding author.

## Acknowledgements

We thank Andre Kiesel for critical revision of this manuscript. This work was supported by the National Science and Technology Major Project (No. 2018ZX10101002-001-001), National Key Research and Development Program Projects of China (2017YFD0500305), the State Key Program of National Natural Science of China (U1808202), NSFC International (regional) cooperation and exchange program (31961143024), the Key-Area Research and Development Program of Guangdong Province (2018B020241002), and the Guangdong Provincial Science and Technology Project (2018B020207013).

## Authors’contribution

Z. Chen and J. Lu conceived and directed the study, Q. Zhang performed the temporal and spatial distribution analysis, Y. Lu, Z. Guo, W. Zhang, C. Guo participated in discussion and improvement of the contents, Q. Zhang and X. Zhang participated in figure and table preparation. C. Liao, X. Han and Q. Li collected the case data. Y. Lu performed critical revision of the manuscript. Z. Chen wrote the paper with help from all others.

